# Role of heterogeneous infection histories on variant dynamics: exact and approximate approaches

**DOI:** 10.64898/2026.10.03.26364599

**Authors:** Pourya Toranj Simin, Pierre-yves Boëlle, Chiara Poletto

## Abstract

Host immunity acquired through prior infections is a key factor shaping the spread of emerging variants. However, accounting for complete infection histories in mathematical models is challenging because the number of possible histories grows exponentially with the number of variants. Here, we present a flexible framework for modeling the co-circulation of multiple variants emerging at arbitrary times while incorporating cross-immunity and variant-specific characteristics. We introduce three tractable approximations: *truncation*, which retains only infection histories up to a given number of strains; *one variant at a time*, which is exact when variants circulation does not overlap; and *two variants at a time*, which is exact when at most two strains co-circulate. We systematically evaluate these approximations across a range of cross-immunity levels and inter-variant emergence delays. *Truncation* approximations perform best under short emergence delays and strong cross-immunity, whereas the other approaches outperform them at longer delays. When applied to SARS-CoV-2 variant dynamics, the second-order truncation and the *two variants at a time* approximation accurately capture incidence patterns, whereas the other approaches perform well only for specific subsets of variants. These results identify the parameter regimes in which simplifying assumptions remain valid, provide guidance for model selection, and highlight the key role of infection history in shaping epidemic trajectories.

## Introduction

Many human viruses, including influenza, SARS-CoV-2, and dengue, circulate as phenotypically distinct strains, making strain structure a central feature of pathogen dynamics [1–3]. When a new strain emerges, it may replace resident strains or coexist with them, shaping pathogen evolution [2]. These dynamics have important implications for infection risk, vaccine design, and the emergence of drug resistance [4–6]. A large body of work in infectious disease ecology has examined how pathogen traits, mechanisms of interaction among strains, and host characteristics jointly shape variant circulation and evolutionary outcomes over time [7–17].

The COVID-19 pandemic illustrates the consequences of the emergence of multiple variants of concern and the rapid adaptation of SARS-CoV-2 to the human population [3, 14, 16]. The appearance of new variants has repeatedly triggered sudden surges in cases, hospitalizations, and deaths, with major implications for epidemic assessment and response planning [18–22]. These events highlight the need to model multi-variant interactions not only to understand evolutionary dynamics but also to support real-time outbreak analysis [3, 23, 24], and call for a better understanding of the factors shaping incidence dynamics following variant emergence.

Changes in incidence depend in complex ways on viral characteristics—antigenicity, transmissibility, and time course of infectivity—as well as on host factors such as behavior and pre-existing immunity [7–16, 23]. In particular, the host immune response is a central factor mediating interactions among multiple strains: immunity induced by infection with one strain may confer partial protection against others, so that an individual’s susceptibility to a circulating variant depends on their infection history. Explicitly representing this dependence in population-level dynamics typically requires tracking an exponentially growing number of immune states [1, 10, 25]. To address this challenge, several studies have developed reduced mathematical frameworks that characterize how immune histories shape endemic equilibria of multi-strain systems, including conditions for strain coexistence [1, 10, 25, 26]. By contrast, limited theoretical work has focused on the description of out-of-equilibrium multi-strain incidence dynamics [27, 28].

Nevertheless, empirical patterns may justify simplifying assumptions. Strong cross-protection between variants may limit the diversity of infection histories that contribute substantially to transmission. In addition, long intervals between successive variant emergences may limit temporal overlap in their circulation, and new variants often dominate for a period before being replaced. Models could thus focus on the few variants circulating at any given time while retaining the effects of past infections on population immunity. Together, these considerations motivate tractable approximations for describing variant dynamics in pathogens such as SARS-CoV-2 and influenza viruses.

Here, we derive equations describing variant dynamics in the general setting of partial immune evasion. The resulting framework applies to an arbitrary number of co-circulating strains and can accommodate strain-specific transmissibility and variable cross-immunity. By representing infection history as an unordered set of previously encountered strains, we show that tractable approximations of the full equations are possible in certain cases, which allow more efficient numerical simulations of many-strain systems. We systematically quantify the effectivness of these approximations, thus providing insights on the role of population immunity on the multistrain dynamics.

## Methods

### Renewal equation for interacting strains

#### Model setting and assumptions

We adapt the renewal-equation formalism to the co-circulation of multiple interacting strains [29]. A key advantage of this approach is that it explicitly accounts for the full generation-time distribution— not only its mean—which mediates the relationship between the basic reproduction number and observed incidence growth. Because generation times may vary across variants and with host immune status, properly accounting for this variation (via renewal equations rather than rate equations) is important for characterizing variant growth dynamics [30, 31].

For a single virus in a closed population, the renewal equation links incidence at time *t, I*(*t*), to past incidence *I*(*t − τ*) as 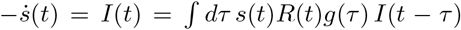, where *s*(*t*) is population susceptibility, *R*(*t*) is instantaneous reproduction number (that can change over time due to interventions, behaviours or environmental drivers), and *g*(*τ*) is the generation-time distribution. We write *β*(*t, τ*) = *s*(*t*)*R*(*t*)*g*(*τ*) to simplify equations. To generalize this equation to the context of multiple strains, we assume that (1) a host can only be infected with one strain at a time, (2) there may be cross-immunity between strains, and (3) immune response does not change with the order of infections.

To model cross-immunity, we adopt a history-based approach, whereby hosts are labeled according to their history of past infections [9, 10, 32]. This infection history affects both susceptibility to a given strain and infectiousness when infected by that strain. We assume that infection order has no effect on subsequent immunity – we thus ignore immune imprinting effects [33, 34]. Infection history *h* is thus represented by the unordered set of strains previously encountered by an individual, starting with *h* = *∅* for those who have never been infected before. Under this assumption, the number of possible immune histories grows exponentially with the number of *n* strains.

We define:

- 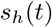 as the proportion of individuals with infection history *h* who, at time *t*, are not infected;
- 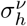 as the susceptibility to variant *v* of individuals with infection history *h*, ranging between 0 (full protection against *v*) and 1 (no protection against *v*). Without loss of generality, we set 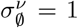 for all variants, and allow homologous reinfection if 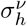 for *v ∈ h* is not set to 0. In this case, however, the modeled immunity to *v* does not depend on the number of times *v* has been encountered in the past, but for the first time;
- 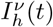 as the incidence of variant *v* at time *t* among individuals with infection history *h*;
- 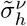 as the relative infectiousness of *v* infected individuals depending on infection history *h*, ranging between 0 (not infectious) and 1 (fully infectious). Here again, we set, without loss of generality, 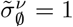 for all variants;
- 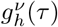 as the generation time distribution in those infected with *v* with history *h*, to account for differences across variants and immune histories [35, 36].
- 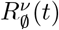 as the basic reproduction number of variant *v* in a fully naive population. This quantity is indexed by time to account for change in transmission conditions affecting *β*, as discussed above. Variants may differ in their epidemiological traits. In practice, the effective reproduction number of variant *v* will depend on the immune profiles of hosts.

#### Renewal equations

Under the assumptions above, the renewal equation for a multi-strain system can be written as

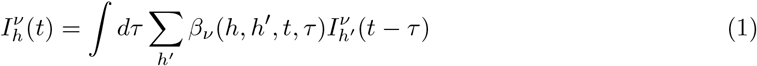

with *β*_*v*_ (*h, h*^*′*^, *t, τ*), being the infectiousness, which we decompose as:

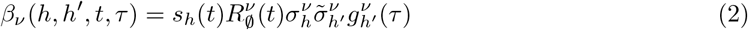

Substituting Eq. 2 into Eq. 1, the later can be rewritten into the following terms:

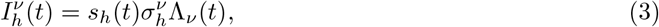

where Λ_*v*_ (*t*) is the force of infection. Eqs. 1 to 3 describe the transmission dynamics in a closed population. We can incorporate variant introducitons by adding to Λ_*v*_ (*t*) an importantion term *F*_*v*_ (*t*), which can be time-varying and variant-specific. The complete expression of Λ_*v*_ (*t*) is thus

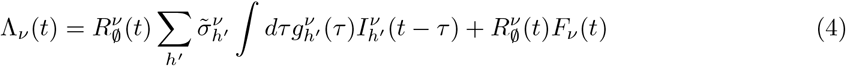

To compute the change in the susceptible population, 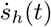, with infection history *h*, we need to account for individuals leaving *s*_*h*_(*t*) as they become infected, individuals entering *s*_*h*_(*t*) as they recover from an infection with variant *u* and prior infection history *h −{u}*, 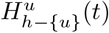, and individuals re-entering *s*_*h*_(*t*) as they recover from a homologous reinfection with variant *v* and prior infection history *h* with 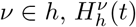:

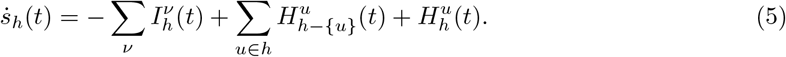

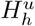 can be computed from the incidence, by knowing the protection-duration distribution 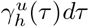, which a priory depends on the variant *v* and the history of infection *h*:

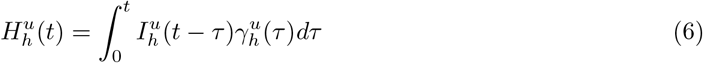

Importantly, 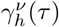 need not equal 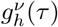. The former describes the timing an infividual become susceptible again after infection, whereas the latter describes the timing of transmission. Individuals may cease transmitting before becoming susceptible again because of hospitalization, isolation, or temporary protection against reinfection.

#### Computing quantities

Eqs. 3, 5, and 6 together describe the dynamical system of circulating strains. Other quantities can then be derived. For instance, from 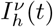 we can compute 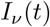, the total incidence of variant *v*, as follows:

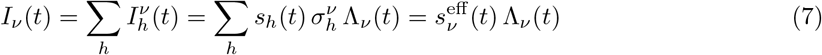

where we define the effective susceptibility to variant *v* as

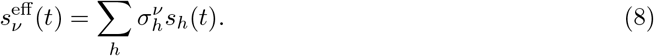

Similarly, we can define and compute the total incidence of all variants as

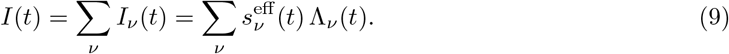

Finally, we compute the instantaneous reproduction number of each variant, *R*^*v*^ (*t*). For a singlevirus renewal equation, it is obtained by the incidence equation and reads *R*(*t*) = ∫*β*(*t, τ*) *dτ* [37]. Extending the approach to the multi-variant case (see Supplementary Information) gives

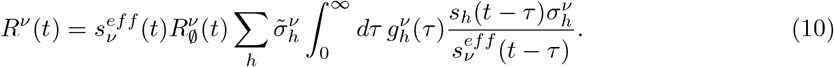

If the generation-time distribution is independent of infection history, 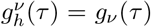, and infection history does not reduce infectiousness, 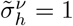, this expression simplifies to

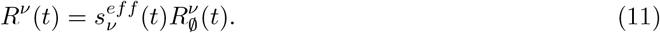

#### Specifying cross-immunity

The specification of the cross-immunity parameters 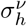, is challenging because, although pairwise cross-immunity may be estimated from data, extending these estimates to hosts with multiple previous infections requires an assumption about how immune effects accumulate [1].

One commonly used approach is the *product* cross-immunity formulation [38], in which reductions in susceptibility or transmissibility accumulate multiplicatively. Under this assumption, the crossimmunity against variant *v* due to infection history *h* is given by 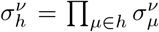, where 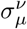 is the cross-immunity between variants *µ* and *v*.

An alternative approach is the *minimum* cross-immunity formulation [38], in which susceptibility to variant *v* is reduced to the strongest (i.e., smallest) reduction factor among all previously encountered variants, i.e., 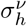 = min 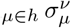. In both cases, protection against a circulating variant tends to increase with the number of past infections, i.e., 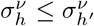 if *h*^*′*^ *⊂ h*. In the *product* cross-immunity formulation, 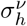 diminishes rapidly as the size of *h* increases, unless all pairwise cross-immunity are weak (i.e., 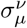 is close to 1 for all *v* and *µ*). In the *minimum* cross-immunity formulation, the protection against a given variant conferred by a set of previous infections is dominated by the most closely matching variant encountered in the past.

Note that, similar considerations as above apply also to the term for the reduction in infectiousness, 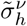.

### Solving the equations

The equations introduced in the previous section can be solved numerically, although the number of equations grows exponentially with the number of variants. Below, we describe some ways computations can be simplified and the corresponding regimes.

#### Truncating long histories

The *truncation* approximation corresponds to situations in which long infection histories are rare, for example following initial emergence, or, practically, if cross-immunity tends to decrease susceptibility to infection in those with several past infections, limiting their role in the overall transmission dynamics. To highlight the role of different infection histories, we can rewrite Eq. 4 as follows:

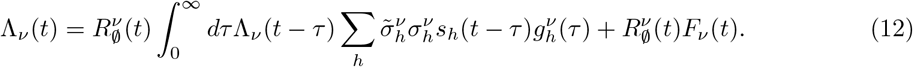

If 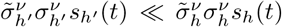 whenever *h ⊂ h*^*′*^, and provided 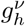 does not vary substantially with *h*, then terms of Eq. 12 corresponding to longer immune histories contribute little to the force of infection. In this case, we can truncate Eq. 12 by retaining only histories up to cardinal *m*. The resulting system consists of 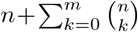 coupled integro-differential equations, so the computational complexity grows polynomially with *m, O*(*n*^*m*^).

For example, for three variants (*v* = 1, 2, 3), the sets corresponding to the zero-, first-, and second-order truncation are

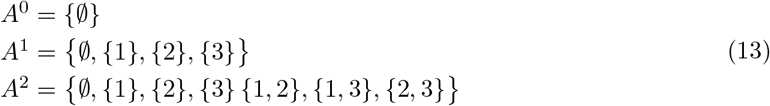

Further details of the approximation method are provided in the Supplementary Information.

#### One variant at a time

In the *one-variant-at-a-time* approximation, we assume that each variant emerges only after the epidemic caused by its predecessor has ended. The problem therefore reduces to the spread of a single variant in a population stratified by infection history, with the size of each history class *h* determined by previous variant epidemics. When variant *c* emerges at time 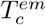, infection histories can contain only variants that circulated before *c*. Because 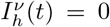 for all *v* ≠ *c* after 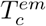, population immunity to previous variants remains fixed. Partitioning infection histories according to whether they contain *c*, and assuming that individuals who recover from variant *c* cannot be reinfected by it, i.e., 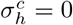 for *c ∈ h*, Eq. 5 simplifies to

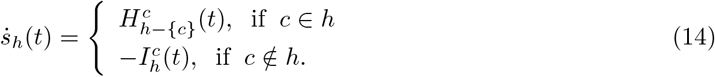

Thus, histories containing *c* are populated through infections of individuals with histories that do not yet contain *c*, whereas histories that do not contain *c* are depleted as individuals become infected with *c*. The resulting dynamics can be reduced to a self-consistent equation for the force of infection, Λ_*c*_(*t*) (see Supplementary Information for the derivation), which can be solved separately for each emerging variant:

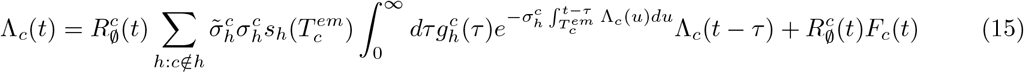

The resulting distribution of infection histories at the end of the epidemic caused by variant *c* provides the initial condition for the subsequent emerging variant *d*, yielding a stepwise description of the multi-strain dynamics. Since the approximation assumes that only one variant circulates at a time, it is defined only up to the emergence of the next variant, which restricts the domain over which the approximation is defined. The method is exact when epidemics caused by successive variants do not overlap and is used as an approximation when they do. Its accuracy under overlapping epidemics is evaluated in the Results section.

#### Two variants at a time

We last introduce the *two variants at a time* truncation as an extension of the previous approach, allowing for the co-circulation of at most two variants. This allows partial overlap between epidemic waves while remaining computationally tractable. Consider variant *c*, which emerges at time 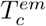 while the previously emerged variant *b* is still circulating. During the interval 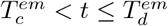, where *d* denotes the next emerging variant, we assume that only *b* and *c* contribute to transmission, i.e.,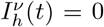 for all *v* ≠ *b, c*. By assuming that individuals cannot be reinfected by the same variant and following the same logic as in *one variant at a time*, we partition the set of infection histories *h* according to whether they contain variants *b* and/or *c*. Eq. 5 can then be written as:

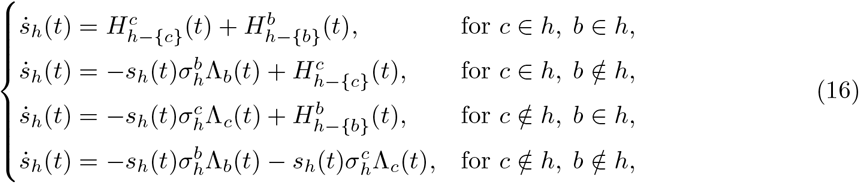

where the forces of infection of the two circulating variants, *v* = *b, c*, are given by:

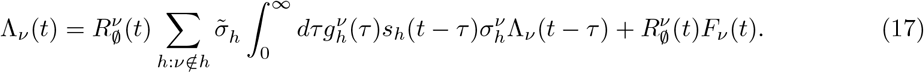

Additional details of the derivation of the equations are reported in the Supplementary Information. Unlike the *one variant at a time* framework, the two forces of infection are mutually coupled through the shared susceptible population and therefore cannot generally be reduced to a single closed-form equation. The dynamics nevertheless depend only on the current pair of circulating variants and the distribution of infection histories at 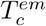. The resulting system is solved iteratively over successive intervals, using the infection-history distribution at the end of each interval as the initial condition for the next. The accuracy of this approximation when more than two variants co-circulate is evaluated in the Results section.

### Numerical experiments

We examine the model dynamics to evaluate when the three approximations introduced above can be accurate. We compare the resulting variant-specific incidence with the exact and approximated solutions and compute relative or absolute errors, as appropriate, under two different model settings. Abolute error was defined as the difference between the approximate and exact incidence curves, while relative error was obtained by diving the absolute error by the exact incidence.

#### Role of cross-immunity and timing of variant emergence

We first examine how cross-immunity and the interval between variant introductions affect approximation accuracy in a system of four variants. We assume that infection history does not affect infectiousness 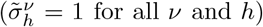, while susceptibility is rescaled uniformly such that 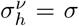 if *v ∈/ h* and 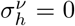 otherwise, thereby preventing homologous reinfection. The variants emerge at emergence times, 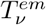, equal to 0, Δ*t*, 2Δ*t*, and 3Δ*t*, with the first representing the ancestral variant. We systematically vary Δ*t* and *σ*. To isolate their effects, we keep transmission and natural-history parameters identical across variants and infection histories, so that 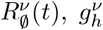, and 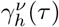 do not depend on *v* or *h*. We set 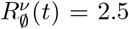 and assume that both the generation-time and protection-duration distributions follow gamma distributions with a mean of 5 days and a standard deviation of 2 days. For the initial seeding, we assume that all imported cases are newly infected individuals with no prior infection history. These cases are introduced at 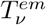, with the initial number of infected individuals 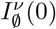 of the population. Accordingly, for 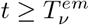, the importation term of the force of infection takes the form 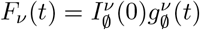.

#### SARS-CoV-2-like scenario

To exemplify the approximations in a realistic example, we consider the initial seasons (2020 to 2022) following emergence of SARS-CoV-2, where the historical lineage was followed by the Alpha, Beta, Delta, and Omicron variants in France.

We calibrated the model to reproduce the observed co-circulation pattern, accounting for variant emergence times, variant-specific epidemiological characteristics, and pairwise cross-immunity estimates from the literature. SARS-CoV-2 incidence and variant frequency is reported in the Supplementary Information, Figure S1. We assumed that infection history does not reduce infectiousness, 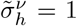, and derived cross-immunity for each infection history from the pairwise parameters using the minimum rule. The generation-time distribution, *g*(*τ*), and protection-duration distribution, *γ*(*τ*), were identical across variants and infection histories and followed gamma distributions with means of 5 and 28 days and standard deviations of 2 and 14 days, respectively, consistent with COVID-19 estimates [39–41]. Finally, we used temporal changes in the stringency index to modulate the reproduction number of each variant as 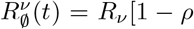 Stringency(*t − t*_delay_)]. Data on the stringency index where obtained by [42]. The parameters *ρ* and *t*_delay_ were calibrated against empirical estimates of the effective reproduction number in France obtained from [43]. Calibration was performed over the initial period from 2020/03/01 to 2020/09/01, when the depletion of susceptible individuals and the change in tranmission due to emerging variants were limited. We obtained *ρ* = 0.908 and *t*_delay_ = 19 days. During the simulations we ensured that incidence and population immunity at each variant’s emergence were comparable to COVID-19 values.

We use the SARS-CoV-2-like scenario as an illustrative example with several variants occuring sequentially in time and do not aim to reproduce the French epidemic in detail or to precisely match incidence values. Accordingly, we neglect vaccination and behavioral changes other than those captured by interventions, which are not relevant for the theoretical framework developed here.

## Results

### Goodness of the approximations: role of cross-immunity and timing of variant emergence

Figure 1 shows variant-specific incidence for different values of Δ*t* at *σ* = 0.5. Each panel compares the exact solution (black) with truncation approximations of different orders (shades of yellow), the *one variant at a time* approximation (light purple), and the *two variants at a time* approximation (dark purple). As expected, the *truncation* error decreases as higher-order terms are retained. However, the accuracy of a given truncation order depends on both the variant considered and the delay between successive variant emergences. All truncation orders reproduce the dynamics of the ancestral variant well, but accuracy diminishes for later-emerging variants. While the accuracy for the ancestral variant improves as Δ*t* increases, the opposite trend is observed for emerging variants. Conversely, the *one variant at a time* and *two variants at a time* approximations become more accurate as emergence delays increase. As described in the Methods section, the *one variant at a time* approximation is exact when epidemic waves do not overlap, as for variants 1 and 2 at Δ*t* = 100. The *two variants at a time* approximation is exact when no more than two waves overlap, as for variants 1 and 2 at Δ*t* = 50 and for all variants at Δ*t* = 100. Nevertheless, Figure 1 shows that both approximations can remain reasonably accurate even when their overlap requirements are temporarily violated. For example, the growth of variant 3 at Δ*t* = 100 under the *one variant at a time* approximation closely follows the exact curve despite its overlap with the peak and decline of variant 2. Note that the accuracy of the approximations can also be assessed through the time-varying reproduction number, *R*_*t*_. In Figure S2, we compare *R*_*t*_ obtained from the exact solution and the five approximation methods for Δ*t* = 50 taken as an example.

**Figure 1:**
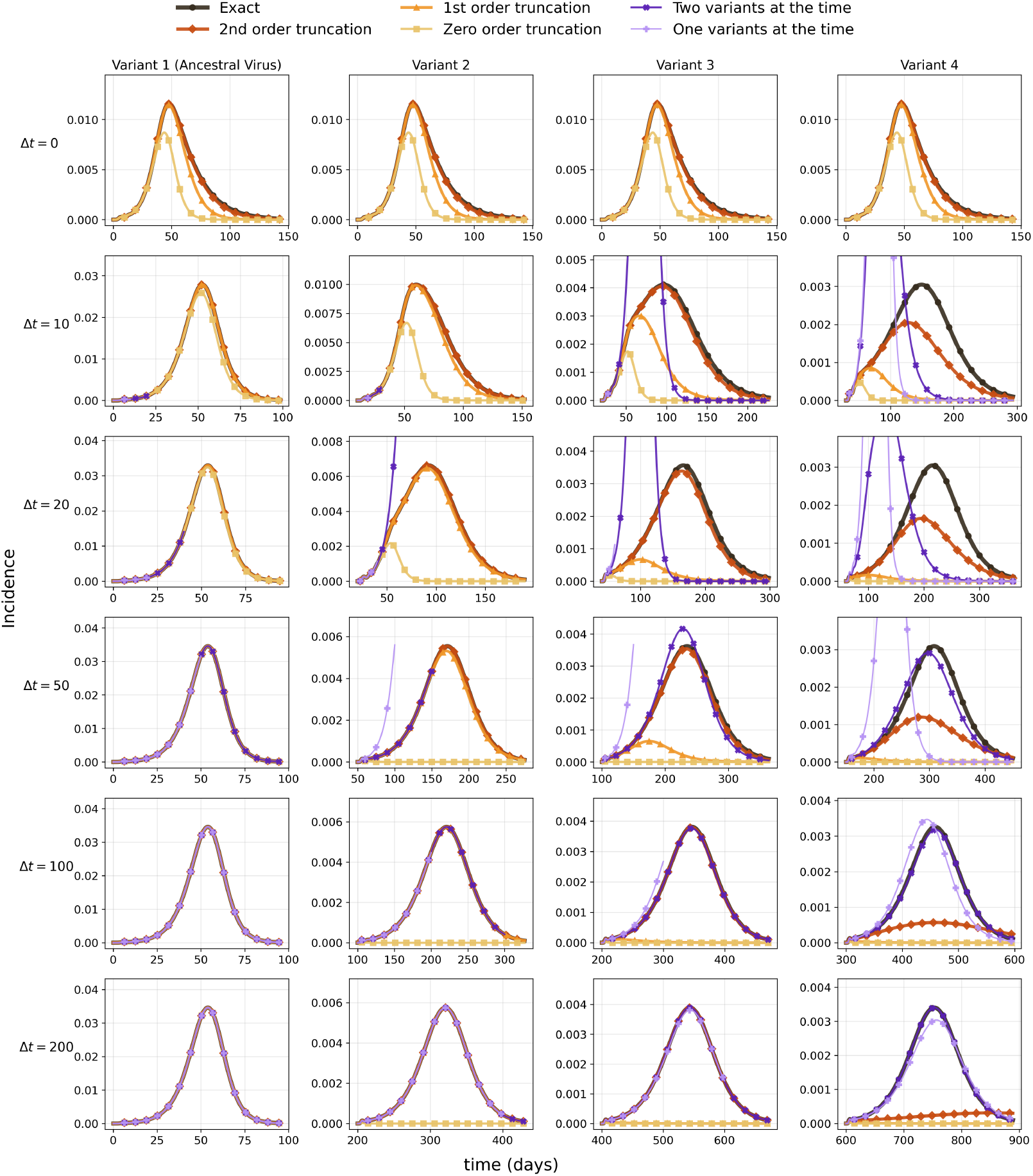
Variant incidence: comparison of the exact model and approximations. Each panel shows incidence as a function of time for each variant (column) and for different emergence delays Δ*t*. Each row corresponds to a different value of the emergence delay. Different line styles represent the calculation methods—exact or approximate. *One variant at a time* is defined only until the next variant emerges, which limits the range of definition of the approximation. Fixed parameters are: *σ* = 0.5, 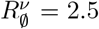, the generation-time *g*(*τ*) and recovery-time *γ*(*τ*) distributions are Gamma distributions with mean 5 days and standard deviation 2 days.

These results were obtained for a fixed value of *σ*. To systematically explore different values of *σ* and Δ*t*, we define a synthetic indicator of approximation accuracy and plot it as a function of both parameters. We focused on the growth phase of each variant and computed the relative error integrated from the start up to the peak of the exact curve. In Figure 2, different rows correspond to different approximations and different columns to different variants.

**Figure 2:**
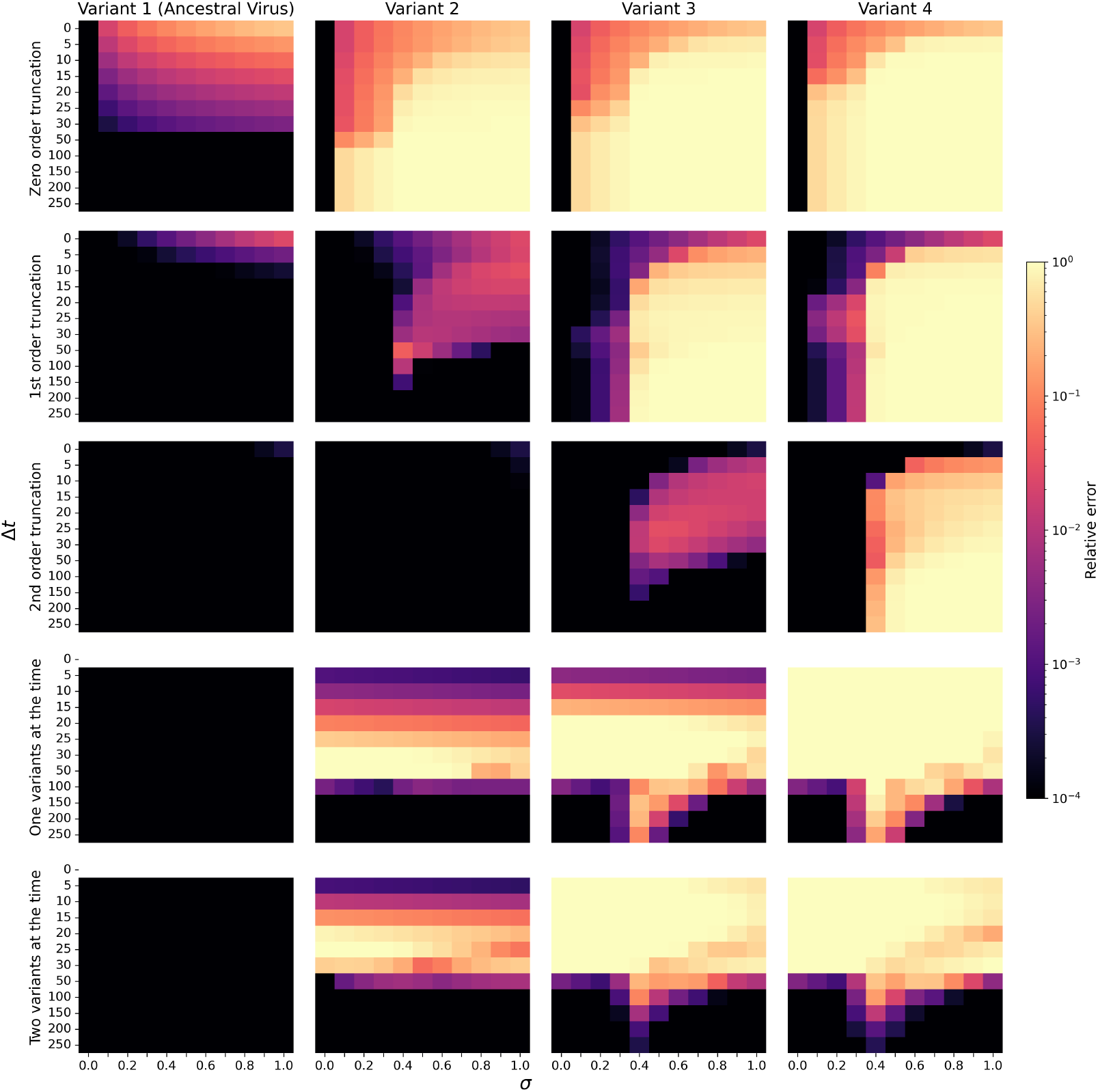
Relative error of the different approximation methods. Panels show the relative error of each approximation for each variant as a function of *σ* and Δ*t*. The relative error is computed over the incidence up to the peak. Rows correspond to the approximation methods, while columns correspond to the four variants. The color scale shows the relative error of the incidence. The scale is saturated for relative errors below 10^*−*4^ and above 1 to highlight the region corresponding to intermediate relative errors. The *one variant at a time* and *two variants at a time* approximations are not defined for Δ*t* = 0; consequently, the corresponding cells in the heat maps are left blank.

The heatmaps in Figure 2 clarify the patterns already apparent in Figure 1. All truncation orders accurately reproduce the incidence of the ancestral variant, with a maximum relative error of 13% for the zero-order truncation when all variants emerge simultaneously. As Δ*t* increases, interference from emerging variants weakens and the error decreases. Relative errors are generally higher for emerging variants. For the *v*-th variant, truncation orders *m ≥ v −* 1 yield relative errors below 3%. At the time of its emergence, individuals can have encountered at most the *v −* 1 preceding variants, and these infection histories are retained by such truncations. The accuracy of the (*v −* 1)-order truncation indicates that longer infection histories formed during the growth of variant *v* do not substantially affect its dynamics. By contrast, when *m < v −* 1, the relative error is higher and increases with both Δ*t* and *σ*. Increasing Δ*t* allows longer infection histories to accumulate, while increasing *σ* (i.e., weakening cross-immunity) promotes reinfection; both effects increase the prevalence of histories omitted by the truncation and reduce its accuracy. The error remains below 10% for Δ*t ≤* 5 across almost all values of *σ*, whereas for larger Δ*t* this occurs only when *σ* is below approximately 0.3–0.4. For these values of *σ* the emerging variants do not spread widely as *σ ≃* 0.35 corresponds to the emergence threshold above which, the emerging variant has an effective reproduction number greater than 1 and can invade the population (Supplementary Information, Figure S3).

For the *one variant at a time* and *two variants at a time* approximations, the error approaches zero for large Δ*t* (Figure 2). At small Δ*t*, however, the error pattern varies among variants. For variant 2 under both approximations, and for variant 3 under the *one variant at a time* approximation, the error is largest when the variant emerges near the incidence peak of the ancestral variant, because the cumulative incidence of the ancestral variant is then increasing rapidly, a change neglected by the approximations. The effect of *σ* also depends on Δ*t*: for short emergence delays, the error tends to decrease as *σ* increases (the opposite trend to that observed for the truncation approximations), whereas for longer delays it peaks near the emergence threshold. In this parameter region, the dynamics are slower and co-circulation is prolonged.

To compare the approximations, we identify, for each variant and each (*σ*, Δ*t*) pair, which of the five approximations yields the minimum error. When determining the minimum, we treat all errors below 10^*−*4^ as equivalent because their differences are not meaningful. Figure 3 shows that incidence up to the epidemic peak can be described with an error below 10^*−*4^ for the ancestral variant and variant 2. For variant 3, the maximum error is of order 10^*−*2^ and is restricted to a limited parameter region, whereas for variant 4 it reaches 10^*−*1^ over a wider region. Truncation approximations generally perform best, particularly when Δ*t* is not large. For all variants except variant 4, truncation remains among the best-performing approaches at high Δ*t*, alongside either the *one variant at a time* or *two variants at a time* approximation. The second-order truncation is the best-performing truncation in most cases. In some regions, however, it provides no meaningful advantage over the first-order truncation (variant 2 at high Δ*t*) or even the zero-order truncation (some parameter regions for the ancestral variant). For variant 4, both the *one variant at a time* and *two variants at a time* approximations substantially outperform truncation at high Δ*t* and perform equivalently at the largest values of Δ*t*.

**Figure 3:**
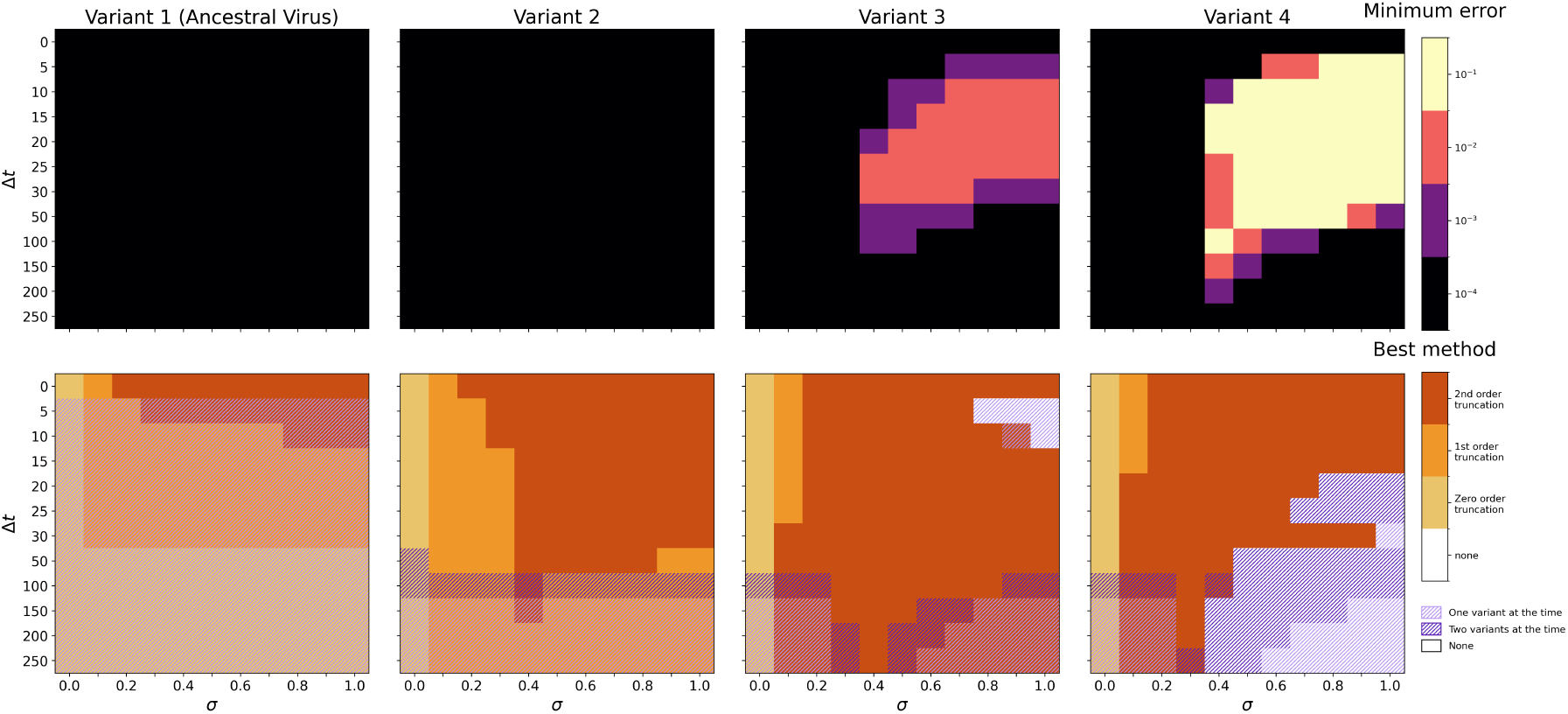
Top row: minimum relative error obtained among the five approximations considered. Bottom row: approximation yielding the minimum error. When a truncation approximation is indicated as the best-performing approach, all higher-order truncations perform equivalently. Similarly, when the *one variant at a time* approximation is indicated, the *two variants at a time* approximation performs equivalently. Errors below 10^*−*4^ are treated as equivalent. Parameters are as in Figure 2.

### Goodness of approximation: SARS-CoV-2-like scenario

To better explore the effect of the delay of variant emergence and the role of cross-immunity in a heterogenous case, we now consider a scenario which mimicks the co-circulation of the ancestral SARS-CoV-2 strain and the Alpha, Beta, Delta, and Omicron variants in France between 2020 and 2022. The emergence times, the epidemiological characteristics, and the cross-immunity interactions among variants taken from the literature are indicated in Table 1. Figure 4a displays the crossimmunity matrix 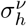 for each variant and possible infection history.

**Table 1:** Parameters of the COVID-19 scenario in France.

| Variant | Basic<br>Reproduction<br>Number | Emerging<br>time | Immunity evasion with<br>respect to previous<br>variants |
| --- | --- | --- | --- |
| 1: Ancestral virus (Anc v) | $R_0 = 3$ [30] | Day 0 | 0 |
| 2: Alpha | $R_\nu = 1.6R_0$ [44] | Day 255 | 0 w.r. Anc v [44] |
| 3: Beta | $R_\nu = 1.1R_0$ [45] | Day 320 | 0.5 w. r. Alpha and Anc v [44, 46] |
| 4: Delta | $R_\nu = 1.7R_0$ [46] | Day 450 | 0.25 w. r. Beta, Alpha and Anc v [46] |
| 5: Omicron | $R_\nu = 2.1R_0$ [46] | Day 635 | 0.6 w. r. Delta, Beta, Alpha and Anc v [46] |

**Figure 4:**
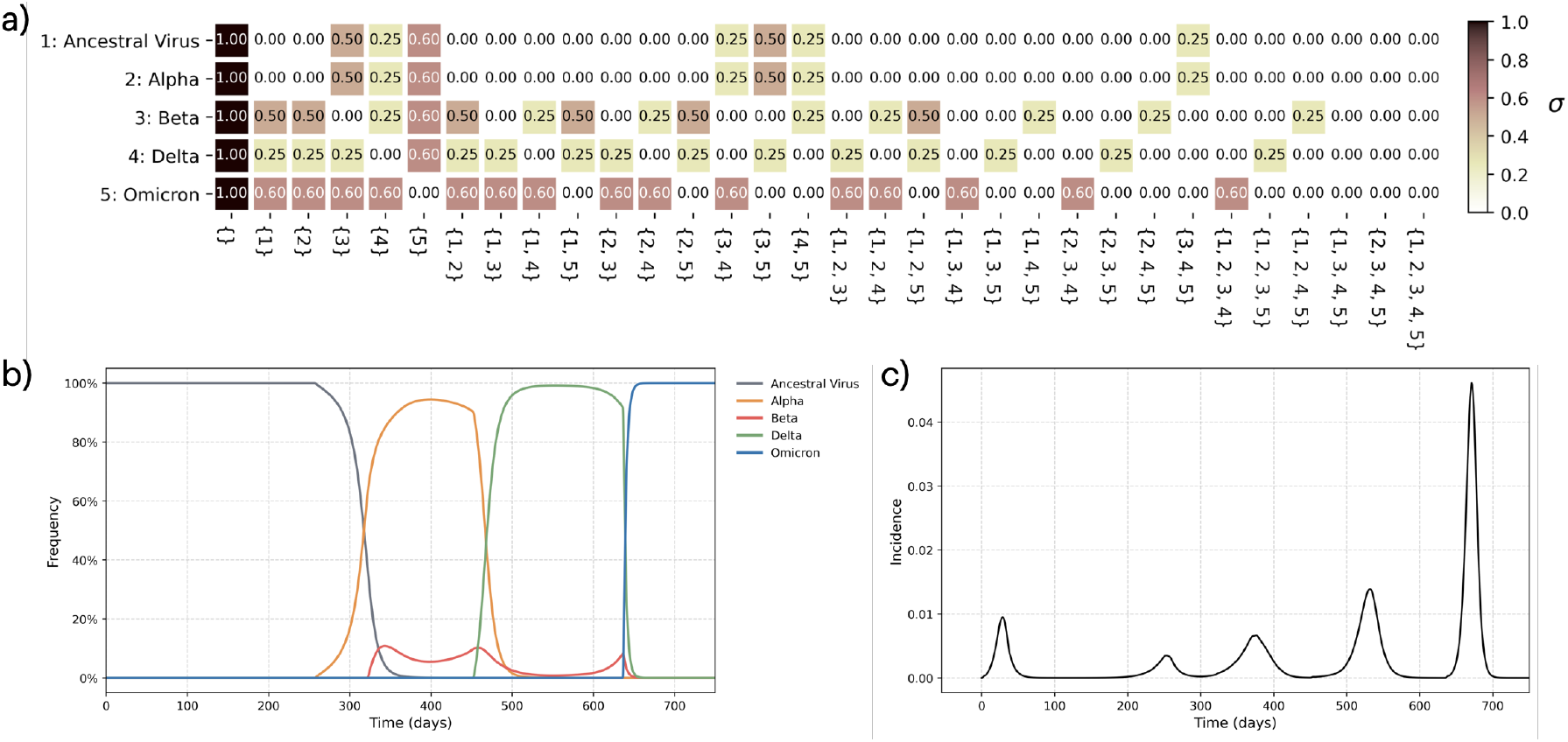
Cross-immunity structure and the corresponding incidence and variant-frequency dynamics for the COVID-19-like scenario obtained from the model. Panel (a) shows the cross-immunity matrix, 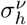, where the rows correspond to viral variants and the columns represent the different combinations of possible infection histories. Panel (b) displays the variant frequencies predicted by the model, defined as the proportion of individuals infected with a given variant relative to the total infected population. Panel (c) shows the total incidence predicted by the model.

We first use the exact model to compute the incidence of each variant and their relative frequencies over time. Figure 4b and c show the succession of epidemic waves caused by the different variants. Although, as discussed above, the model was not fine-tuned to quantitatively reproduce the COVID-19 pandemic in France, the plots qualitatively resemble the incidence and variant-frequency patterns observed for COVID-19 (Supplementary Information, Figure S1). In particular, the emergence of Beta alters the circulation of Alpha, with the two variants coexisting for a limited period until the more transmissible Delta variant emerges and suppresses the previously circulating variants. Subsequently, the Omicron variant undergoes rapid growth, driven by its pronounced immune evasion relative to earlier variants.

We then evaluate the performance of the approximation methods in describing variant cocirculation in this scenario. Figure 5 presents the total incidence with exact and approximate methods (panel a), the absolute errors of each approximation (panel b), and the incidence of each variant over time as computed with exact and approximate methods (panel c). Looking at truncation approximations, we find that both the ancestral variant and the Alpha variant are well described by all approximations; even the zero-order truncation accurately reproduces the exact solution. The good performances for the ancestral variant is consistent with the results of the previous section, where the ancestral variant was always well (or reasonably well) described. Alpha is also well described because of the complete cross-immunity assumed here between Alpha and the ancestral variant 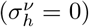. In the example considered in the previous section, strong cross-immunity prevents the variant from emerging, and the resulting subcritical spread is associated with small approximation errors. Here, Alpha can emerge because of its greater transmissibility. Nevertheless, the errors of all truncation approximations remain negligible, indicating that their accuracy is primarily driven by strong cross-immunity, regardless of incidence levels, rather than being limited to cases of subcritical spread.

**Figure 5:**
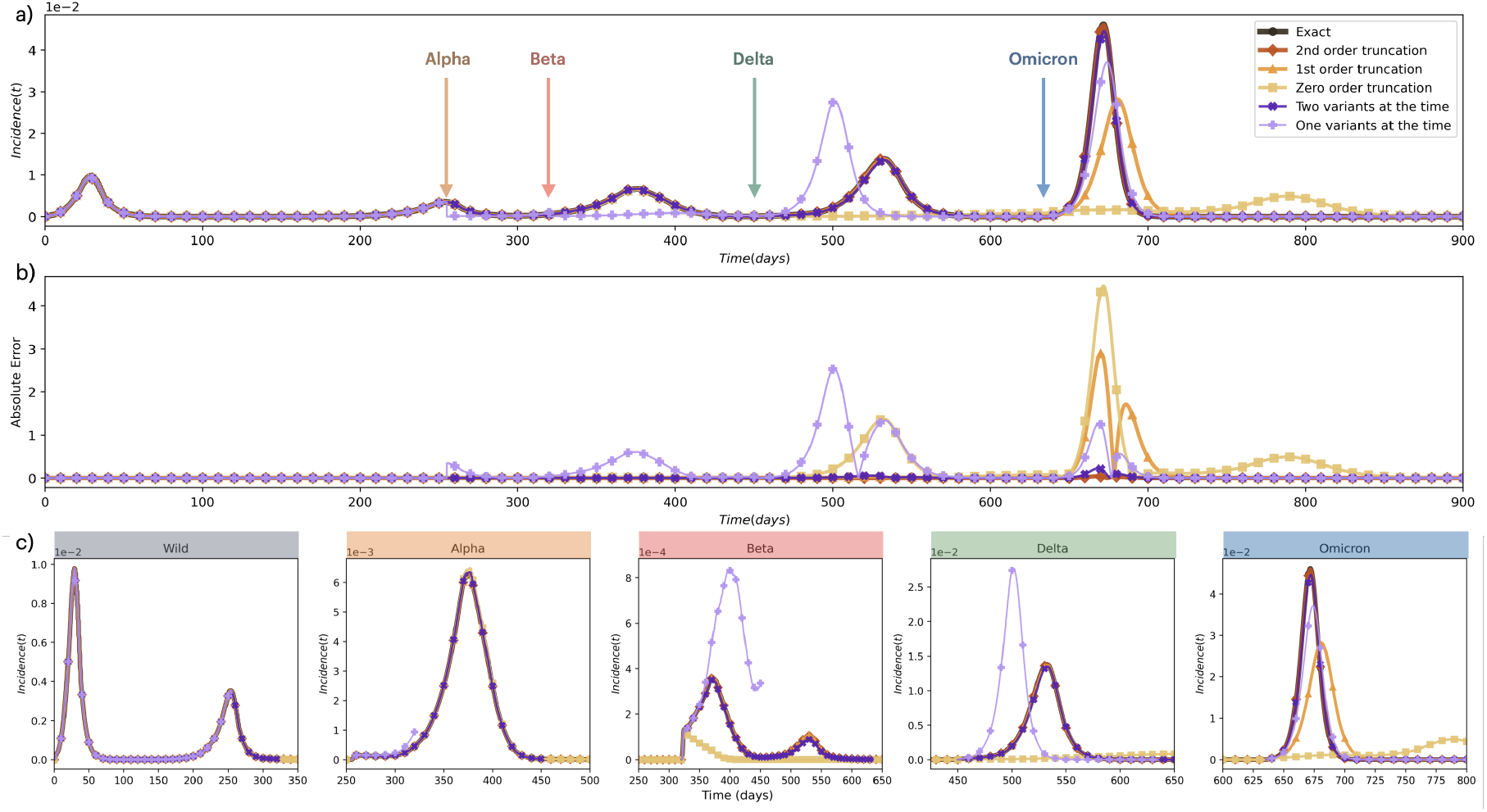
Comparison of the exact solution and different approximations in COVID-19 epidemic scenario. Panel (a) shows the total incidence of infection, with vertical arrows indicating the emergence times of the different variants. Panel (b) presents the absolute error in total incidence for different approximation method. Absolute error is shown instead of relative error because the latter diverges when the exact incidence approaches zero. Panel (c) shows the incidence of each variant over time. The x-axis in all plot indicated time in days. Black curves represent the exact solution and colored curves indicate different approximations.

For the Beta wave and the subsequent Delta wave, the first-order truncation, which retains histories with at most one prior infection, accurately captures the dynamics even though Beta and Delta are the third and fourth variants to emerge in France, respectively. This result suggests that infection histories involving two or more prior infections remain relatively uncommon during these epidemic phases. This is consistent with the results of previous section and can be explained with the immune evasions of Alpha and Delta being absent and low, respectively. In addition, control measures reduce the transmission potential of the ancestral variant below the epidemic threshold, limiting its co-circulation with later variants.

By the time the Omicron wave emerges, however, more complex immune histories have had sufficient time to accumulate due to the partial cross-immunity associated with the Beta and Delta variants. Consequently, higher-order terms must be included in the approximation to accurately capture the Omicron dynamics. Consistent with this interpretation, the first-order truncation becomes insufficient. However, the second-order truncation performs already well and closely matches the exact solution.

The comparison of the *one variant at a time* and *two variants at a time* approximations shows that the former does not adequately reproduce the Beta and Delta waves due to the large overlap between the Alpha and Beta waves, and subsequently between the Beta and Delta waves. Overestimating Delta’s growth has a particularly large effect on incidence because Delta grows faster than the preceding variants. By contrast, the *two variants at a time* approximation is nearly exact over most of the period considered, as the assumption that no more than two variants circulate substantially at the same time is well supported in this setting.

## Discussion

Modeling the emergence of new variants is important for both short- and long-term response planning. A central challenge is capturing the interplay between pathogen antigenicity and host infection history. Previous mathematical work has focused mainly on endemic equilibria [1, 10, 25, 26], whereas the emergence and co-circulation of SARS-CoV-2 variants highlight the importance of transient dynamics in populations with heterogeneous infection histories. Explicitly accounting for the effects of this heterogeneity on incidence growth typically requires a large number of equations. Here, we present a general framework that captures the main factors shaping multi-variant incidence dynamics, including host infection history. We then examine how approximation methods simplify the resulting equations and evaluate their accuracy in both theoretical scenarios and an illustrative epidemic setting inspired by SARS-CoV-2.

The mathematical framework introduced here is general and can accommodate a broad range of epidemiological mechanisms involved in multi-strain circulation. It allows an arbitrary number of variants to emerge at different times and represents host infection history using a history-based approach. Susceptibility, transmissibility, and generation-time distributions can vary with both the infecting variant and the host infection history. By explicitly accounting for time since infection, this formulation extends previous multi-strain approaches that mostly relied on Markovian assumptions [9, 25, 26], in which transition processes such as infection and recovery are memoryless. Because generation-and recovery-time distributions can substantially influence epidemic trajectories and parameter estimates [29–31], relaxing these assumptions provides a more flexible and mechanistically grounded representation of variant emergence. Within this model setting, we derived explicit expressions for variant-specific incidence and the variant-specific time-varying reproduction ratio.

Numerical integration of the full equations becomes cumbersome when more variants are considered. We therefore introduced simplified formulations that substantially reduced computational complexity, justified by the examination of limit cases regarding the emergence of new strains.

First, in the *truncation* framework, we neglected variables corresponding to long immune histories: for a cutoff *m*, only immune histories containing at most *m* previous infections were retained. The rationale is that long immune histories require time to accumulate and could be sufficiently rare to be ignored when new strains occur frequently. In this scenario, susceptible individuals with longer immune histories would also be expected to be more protected against circulating variants, making them less likely to become infected and contribute to transmission. Numerical analysis supported this intuition. The accuracy of *m*-order truncations in general decreases as the interval between successive variant emergences increases—and long immune histories accumulate—and as cross-immunity becomes weaker, which also favors the development of longer immune histories. The approximation accuracy increases with the approximation order and in general the (*v −* 1)-th order truncation well describes the emergence of the *v*-th variant.

Second, in the *one variant at a time* and *two variants at a time* approaches, we simplified the coupling among variants by assuming, respectively, that the number of variants circulating simultaneously is limited to one and two. Under the *one variant at a time* approach, the growth of an emerging variant depended only on the effective susceptibility of the population to that variant. The incidence dynamics of the previously circulating variant can therefore be neglected, provided that its effects on population immunity is retained. When the assumption on variants emerging sequencially holds, the Eq. 15 provides exact results, a result that is interesting in its own right. Similar reasoning has been applied in previous work [47–49]. The spread of an emerging variant on a network can be mathematically described from the properties of the residual network formed by nodes remaining susceptible after the preceding outbreak [47, 48]. More generally, outbreak dynamics can be better interpreted by examining how the susceptibility landscape evolves as the outbreak unfolds [49]. The *two variants at a time* approach extended the same idea by retaining interactions between pairs of co-circulating variants. Also in this case the approach is exact provided only two variants are circualting at the same time. This strategy could be generalized to account for more simultaneously circulating variants, although at the cost of increased computational complexity. We evaluated the accuracy of these approximations when the assumptions about temporal overlap hold only approximately. Our results show that the approximation also performs well when epidemic waves overlap, provided that the overlap is limited in time. As a consequence the approximation accuracy generally increased with the interval between the times of variant emergence. Values of immune evasion close to the emergence threshold - above which the emerging variant has an effective reproduction number greater than 1 even among previously infected individuals - are an exception: in this regime, the error remains high even for long emergence intervals, owing to the particularly slow incidence dynamics around the threshold.

To assess the approximations in a realistic setting, we designed a model that reproduced the key qualitative features of SARS-CoV-2 variant co-circulation in France between 2020 and 2022. We found that the *two variants at a time* approximation reproduced almost all variant waves with high accuracy. The first-order truncation reproduced all waves with good accuracy except the Omicron wave, whereas the second-order truncation also reproduced the Omicron epidemic. The good performance of the second-order truncation suggests that immune histories longer than three infections were not highly prevalent, even though Omicron was the fifth variant to emerge. This pattern can be explained by the strong cross-immunity between Alpha and the ancestral strain and the relatively strong cross-immunity assumed between Delta and all other variants.

Although the model is highly flexible and can accommodate several mechanisms shaping multi-variant dynamics, the analysis presented in the Results section focuses mainly on cross-immunity and the delay between variant emergence times, which play critical roles in emergence dynamics and strongly affect the accuracy of the approximations introduced here. Other mechanisms are not explored in detail, because a broader parameter exploration would be difficult to visualize and discuss. A systematic investigation of how different generation-time distributions influence multivariant dynamics could be pursued in future work. The framework also provides a natural basis for incorporating importation and seeding processes and for examining how their interaction with infection history and cross-immunity shapes epidemic emergence. Finally, stability analyses of endemic equilibria are beyond the scope of the present work but could provide further insight into the behavior of immunologically structured epidemics.

The present version of the model relies on the homogeneous mixing assumption. Population features, such as contact pattern, heterogeneous susceptibility to the infection, and spatial organisation are not taken into account in our framework. These factors are known to influence immune responses and to shape multi-strain dynamics, including change of emergence, growth upon emergence and endemic equilibrium [8, 11, 23, 50, 51]. Still, the renewal equation framework considered here can be directly extended to structured population e.g. by introducing risk classes and risk-stratified transmission rates for each variant.

## Supporting information

Supplementary Table of GISAID Accession Numbers and Genome Metadata

## Data availability

Data on the empirical effective reproduction number in France were obtained from [43] and are available at https://github.com/crondonm/TrackingR/tree/main. Stringency index data were obtained from the Oxford COVID-19 Government Response Tracker (OxCGRT) [42] and are available at https://github.com/OxCGRT/covid-policy-tracker/blob/master/data/timeseries/stringency_index_avg.csv. Daily COVID-19 incidence data for France were obtained from the JHU CSSE COVID-19 Data Repository [52] at https://github.com/CSSEGISandData/COVID-19. Number of SARS-CoV-2 variant sequences in France plotted in the Supplementary Material were obtained from GISAID [53] (https://doi.org/10.55876/gis8.261003md). All code and scripts used for numerical solution, analysis, and visualisation are available at https://github.com/PouryaTS/MultiStrain_RenewalFramework.

## Supporting information

S1 Appendix. Supplementary text and figures. Supplementary Table of GISAID Accession Numbers and Genome Metadata.

## Author contributions

Conceptualization: PTS, PYB, CP. Data curation: PTS. Formal analysis: PTS, PYB, CP. Investigation: PTS, PYB, CP. Methodology: PTS, PYB, CP. Software: PTS. Supervision: PYB, CP. Validation: PYB, CP. Visualization: PTS. Writing – original draft: PTS, PYB, CP. Writing – review & editing: PTS, PYB, CP.

## Acknowledgments

We acknowledge financial support by the Cariparo Foundation through the program Starting Package to CP; Department of Molecular Medicine through the program SID from BIRD funding to CP. We gratefully acknowledge support from the ANRS MIE Network on Modelling Infectious Diseases, which provided a travel grant to PTS for a research visit. The funders had no role in the study design, data collection and analysis, decision to publish, or preparation of the manuscript. We gratefully acknowledge all data contributors, i.e., the Authors and their Originating laboratories responsible for obtaining the specimens, and their Submitting laboratories for generating the genetic sequence and metadata and sharing via the GISAID Initiative, on which this research is based.

