## Supplementary text and figures. for "Role of heterogeneous infection histories on variant dynamics: exact and approximate approaches"

### 1 Supplementary Methods

#### 1.1 Reproduction number

We compute the reproduction number of each variant using the renewal framework. In general, for a renewal equation of the form, $I_\nu(t) = \int \beta(t,\tau)\, I_\nu(t-\tau)\, d\tau$ the corresponding instantaneous reproduction number is defined as $R^\nu(t) = \int \beta(t,\tau)\, d\tau$ [**fraser**][1]. Starting from the renewal equation for $I_h^\nu(t)$, one can derive a renewal equation for the total incidence of variant $\nu$, $I_\nu(t)$, which can be written as:

$$I_\nu(t) = s_\nu^{eff}(t) R_\emptyset^\nu(t) \Big( \sum_h \tilde{\sigma}_h^\nu \int d\tau g_h^\nu(\tau) \frac{s_h(t-\tau)\sigma_h^\nu}{s_\nu^{eff}(t-\tau)}\; I_\nu(t-\tau) + F_\nu(t) \Big) \tag{S1}$$

From this expression, the instantaneous reproduction number $R^\nu(t)$, defined as the expected number of secondary infections generated at time $t$ by an infectious individual of variant $\nu$, is given by

$$R^\nu(t) = s_\nu^{eff}(t) R_\emptyset^\nu(t) \sum_h \tilde{\sigma}_h^\nu \int_0^\infty d\tau g_h^\nu(\tau) \frac{s_h(t-\tau)\sigma_h^\nu}{s_\nu^{eff}(t-\tau)}. \tag{S2}$$

This expression provides a general formulation for the reproduction number of each variant. Under additional simplifying assumptions, it reduces to familiar forms. For example, if the generation-time distribution does not depend on infection history, $g_h^\nu(\tau) = g_\nu(\tau)$, and if infectiousness is not reduced by infection history, $\tilde{\sigma}_h^\nu = 1$, then

$$R^\nu(t) = s_\nu^{eff}(t) R_\emptyset^\nu(t) \int_0^\infty d\tau g_\nu(\tau) \frac{\sum_h s_h(t-\tau)\sigma_h^\nu}{s_\nu^{eff}(t-\tau)} \tag{S3}$$

$$= s_\nu^{eff}(t) R_\emptyset^\nu(t) \int_0^\infty d\tau g_\nu(\tau) = s_\nu^{eff}(t) R_\emptyset^\nu(t) \tag{S4}$$

This shows that the commonly used expression $R^\nu(t) = s_\nu^{eff}(t)\, R_\emptyset^\nu(t)$, holds only under these specific assumptions.

#### 1.2 Solving the equations

We first recall the four equations introduced in the main text that form the basis of the derivations below.

$$I_h^\nu(t) = s_h(t)\sigma_h^\nu \Lambda_\nu(t) \tag{S5}$$

$$\Lambda_\nu(t) = R_\emptyset^\nu(t) \sum_{h'} \tilde{\sigma}_{h'}^\nu \int d\tau g_{h'}^\nu(\tau) I_{h'}^\nu(t-\tau) + R_\emptyset^\nu(t) F_\nu(t) \tag{S6}$$

$$\dot{s}_h(t) = -\sum_\nu I_h^\nu(t) + \sum_{u\in h} H_{h-\{u\}}^u(t) + H_h^u(t). \tag{S7}$$

$$H_h^u(t) = \int_0^t I_h^u(t-\tau)\gamma_h^u(\tau) d\tau \tag{S8}$$

#### Truncating long histories

The *truncation* approximation corresponds to situations in which long infection histories are rare, for example following initial emergence, or, practically, if cross-immunity tends to decrease susceptibility to infection in those with several past infections, limiting their role in the overall transmission dynamics. To highlight the role of different infection histories, we can rewrite Eq. S6 as follows:

$$\Lambda_\nu(t) = R^\nu_\emptyset(t) \int_0^\infty d\tau \Lambda_\nu(t-\tau) \sum_h \tilde{\sigma}^\nu_h \sigma^\nu_h s_h(t-\tau) g^\nu_h(\tau) + R^\nu_\emptyset(t) F_\nu(t). \tag{S9}$$

We can truncate Eq. S4 by retaining only histories up to cardinal $m$. The resulting system consists of $n + \sum_{k=0}^m \binom{n}{k}$ coupled integro-differential equations, so the computational complexity grows polynomially with $m$, $O(n^m)$. For the $m$-order trunctation, we solve the following set of coupled integro-differential equations:

$$\begin{aligned}
&\Lambda^{(m)}_\nu(t) = R^\nu_\emptyset(t) \textstyle\sum_{h\in A^m} \tilde{\sigma}^\nu_h \int_0^\infty d\tau g^\nu_h(\tau) s_h(t-\tau) \sigma^\nu_h \Lambda_\nu(t-\tau) + R^\nu_\emptyset(t) F_\nu(t) \\
&\dot{s}_h(t) = -s_h(t) \textstyle\sum_\nu \sigma^\nu_h \Lambda^{(m)}_\nu(t) + \sum_{u\in h} H^u_{h-\{u\}}(t) + H^u_h(t) \\
&H^u_h(t) = \sigma^u_h \textstyle\int_0^\infty s_h(t-\tau) \Lambda^{(m)}_u(t-\tau) \gamma^u_h(\tau),
\end{aligned} \tag{S10}$$

where $A^m$ denotes the set of all histories with length less than or equal to $m$.

#### One variant at a time

We consider sequential emergence of variants, so that when a new variant emerges, the previous ones have disappeared. At the time $T^{em}_c$ new variant $c$ emerges, the infection histories can only include variants that circulated before $c$; and since all $I^\nu_h(t) = 0$ for $\nu \neq c$ after $T^{em}_c$ immunity against previous strains can no longer change. In these conditions, Eq. S7 simplifies because most terms vanish. Moreover, we can partition the set of histories $h$ into those that include $c$ and those that do not. Eq. S7 can then be written as

$$\dot{s}_h(t) = \begin{cases} H^c_h(t) + H^c_{h-\{c\}}(t), & \text{if } c \in h \\ -I^c_h(t), & \text{if } c \notin h, \end{cases} \tag{S11}$$

which is valid for times $T^{em}_c < t \leq T^{em}_d$ being $d$ the variant emerging after $c$. At this point, we can proceed further by neglecting reinfections with the same variant, i.e., $\sigma^c_h = 0$ for $c \in h$, which implies $H^c_h(t) = 0$, leading to

$$\dot{s}_h(t) = \begin{cases} H^c_{h-\{c\}}(t), & \text{if } c \in h \\ -I^c_h(t), & \text{if } c \notin h. \end{cases} \tag{S12}$$

As immunity against $c$ accumulates, susceptible individuals with infection history $h$, such that $c \in h$, increase. Conversely, susceptible individuals whose infection history does not include $c$ decrease as they become infected with variant $c$. Both terms can be solved explicitly. For histories $h$ such that

$c \in h$, we have

$$\dot{s}_h(t) = \int_{T_c^{em}}^{t} \gamma^c_{h-\{c\}}(\tau)\; I^c_{h-\{c\}}(t-\tau)d\tau = \int_{T_c^{em}}^{t} \gamma^c_{h-\{c\}}(\tau)\; s_{h-\{c\}}(t-\tau)\; \sigma^c_{h-\{c\}}\; \Lambda_c(t-\tau)d\tau; \quad \text{(S13)}$$

while for histories $h$ such that $c \notin h$

$$\dot{s}_h(t) = -s_h(t)\sigma^c_h\Lambda_c(t) \quad \text{(S14)}$$

which gives

$$s_h(t) = s_h(T_c^{em})e^{-\sigma^c_h \int_{T_c^{em}}^{t} \Lambda_c(u)du}. \quad \text{(S15)}$$

Eq. S15 gives the proportion of individuals, among those with history $h$, who are not reached by $c$ at time $t$ during the $c$ epidemic and is computed as one minus the attack rate in the renewal-equation framework for a single varaint epidemic. This expression gives the term $s_{h-\{c\}}(t)$ appearing in Eq. S13, which then becomes:

$$\dot{s}_h(t) = \sigma^c_{h-\{c\}}\; s_h(T_c^{em}) \int_{T_c^{em}}^{t} \gamma^c_{h-\{c\}}(\tau)\; e^{-\sigma^c_{h-\{c\}} \int_{T_c^{em}}^{t-\tau} \Lambda_c(u)du}\; \Lambda_c(t-\tau)d\tau. \quad \text{(S16)}$$

Taken together Eqs. S15 and S16 describe the time evolution of $s_h(t)$ for each $h$ during the course of the $c$ epidemic. Both equations are function of the force of infection of $c$, which can be computed by the following self-consistent equation, obtained by substituing Eq. S15 into Eq. S6:

$$\Lambda_c(t) = R^c_\emptyset(t) \sum_{h:c\notin h} \tilde{\sigma}^c_h \sigma^c_h s_h(T_c^{em}) \int_0^\infty d\tau g^c_h(\tau) e^{-\sigma^c_h \int_{T_c^{em}}^{t-\tau} \Lambda_c(u)du} \Lambda_c(t-\tau) + R^c_\emptyset(t), F_c(t) \quad \text{(S17)}$$

Where the sum runs over infection histories that do not include $c$, because individuals who have recovered from infection with variant $c$ are assumed here not to be reinfected by the same variant, as stated above. Eq. S17 describes the evolution of the force of infection of variant $c$ as susceptibility to $c$ is depleted and immune histories containing $c$ become populated. The assumption of sequential epidemics greatly simplifies the analysis because Eq. S17 can be solved separately for each emerging variant. Since the approximation assumes that only one variant circulate at a time, it is defined only up to the emergence of the next variant, which restricts the domain over which the approximation is defined. The solution gives the distribution of infection histories at the end of each variant epidemic, which can then be used as the initial condition for the next variant $d$, $s_h(T_d^{em})$, yielding a stepwise description of the multi-strain dynamics. The method is exact if the epidemics caused by each variant do not overlap.

##### Two variants at a time

We extend the ne-variant-at-a-time framework to the case in which at most two consecutive variants co-circulate at any given time. Consider variant $c$, which emerges at time $T_c^{em}$ while the previously emerged variant $b$ is still circulating. We assume that all earlier variants have ceased circulating and that no later variant has yet emerged. Thus, $I^\nu_h(t) = 0$ for all $\nu \neq b, c$. During the interval,

$T_c^{em} < t \leq T_d^{em}$, where $d$ denotes the next emerging variant, the dynamics are therefore driven only by variants $b$ and $c$. Following the same approach as for the one-variant-at-a-time approximation, we partition the infection histories according to whether they contain $b$ and $c$. Assuming that individuals cannot be reinfected by the same variant, Eq. S7 becomes

$$\begin{cases} \dot{s}_h(t) = H^c_{h-\{c\}}(t) + H^b_{h-\{b\}}(t), & \text{for } c \in h,\ b \in h, \\ \dot{s}_h(t) = -s_h(t)\sigma_h^b \Lambda_b(t) + H^c_{h-\{c\}}(t), & \text{for } c \in h,\ b \notin h, \\ \dot{s}_h(t) = -s_h(t)\sigma_h^c \Lambda_c(t) + H^b_{h-\{b\}}(t), & \text{for } c \notin h,\ b \in h, \\ \dot{s}_h(t) = -s_h(t)\sigma_h^b \Lambda_b(t) - s_h(t)\sigma_h^c \Lambda_c(t), & \text{for } c \notin h,\ b \notin h. \end{cases} \tag{S18}$$

These equations are valid during the interval $T_c^{em} < t \leq T_d^{em}$, where $d$ denotes the variant emerging after $c$. During this period, variants $b$ and $c$ may circulate simultaneously and both contribute to the infection dynamics. For histories containing both variants, the corresponding susceptible compartment increases as infected individuals recovering from either variant enter this immune-history class. For histories containing $c$ but not $b$, the susceptible population increases through recovery from variant $c$ but decreases because individuals in this class may still be infected by variant $b$. The analogous interpretation applies to histories containing $b$ but not $c$. Finally, for histories containing neither variant, the susceptible population decreases through infections generated by both circulating variants.

The forces of infection of the two circulating variants, $\nu \in b, c$, are given by

$$\Lambda_\nu(t) = R^\nu_\emptyset(t) \sum_{h:\nu \notin h} \tilde{\sigma}_h \int_0^\infty d\tau g_h^\nu(\tau) s_h(t-\tau) \sigma_h^\nu \Lambda_\nu(t-\tau) + R^\nu_\emptyset(t) F_\nu(t). \tag{S19}$$

Unlike the *one variant at a time* approximation, the two forces of infection are mutually coupled through the susceptible population. Each variant therefore affects the population susceptible to the other, preventing the reduction to a single closed-form equation. Nevertheless, variants outside the current pair do not contribute directly to transmission, and the system depends only on the current pair of circulating variants and the distribution of infection histories at $T_c^{em}$. The dynamics can consequently be solved over successive intervals, using the infection-history distribution at the end of each interval as the initial condition for the next.

The *two variants at a time* approximation is exact when no more than two variants circulate simultaneously.

#### 1.3 SARS-CoV-2-like scenario

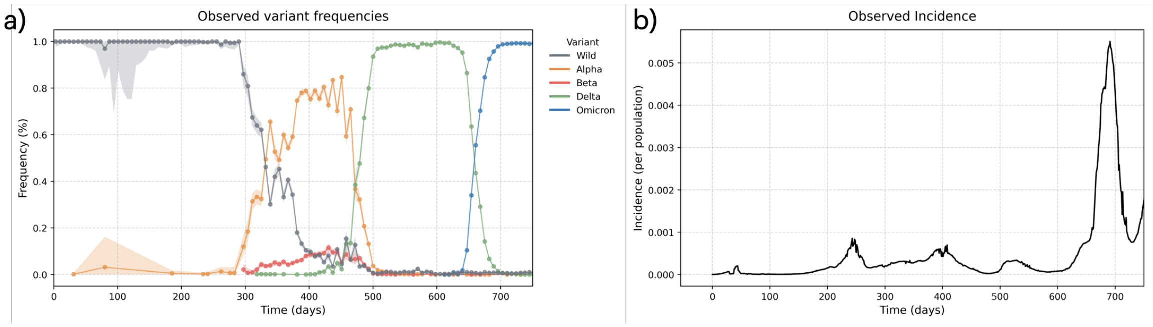

Figure S1: Variant frequency and incidence of SARS-CoV-2 in France used for the calibration of the model in the SARS-CoV-2 like scenario. Panel (a) shows empirical variant frequencies of the historical strain and the Alpha, Beta, Delta, and Omicron variants of concern in France. Number of SARS-CoV-2 variant sequences were obtained from GISAID [2]. Panel (b) shows the empirically observed incidence of SARS-CoV-2 variants in France, obtained from the Johns Hopkins University CSSE COVID-19 Data Repository [3].

### 2 Supplementary Results

#### 2.1 Reproduction number

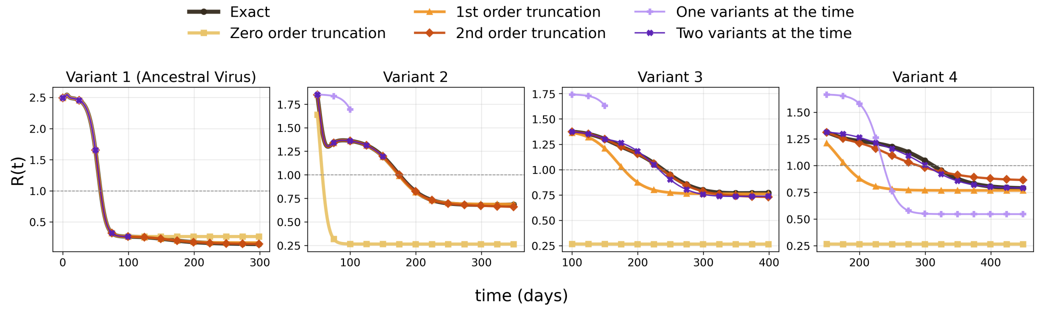

Figure S2: Time-varying reproduction number of the exact model and approximations. Each panel shows the reproduction number as a function of time (days). The first to fourth columns correspond to variants 1 to 4, respectively. Emergence delay and cross-immunity parameter are set to $\Delta t = 50$ and $\sigma = 0.5$ respectivly. All other parameters are as in Figure 1 of the main paper.

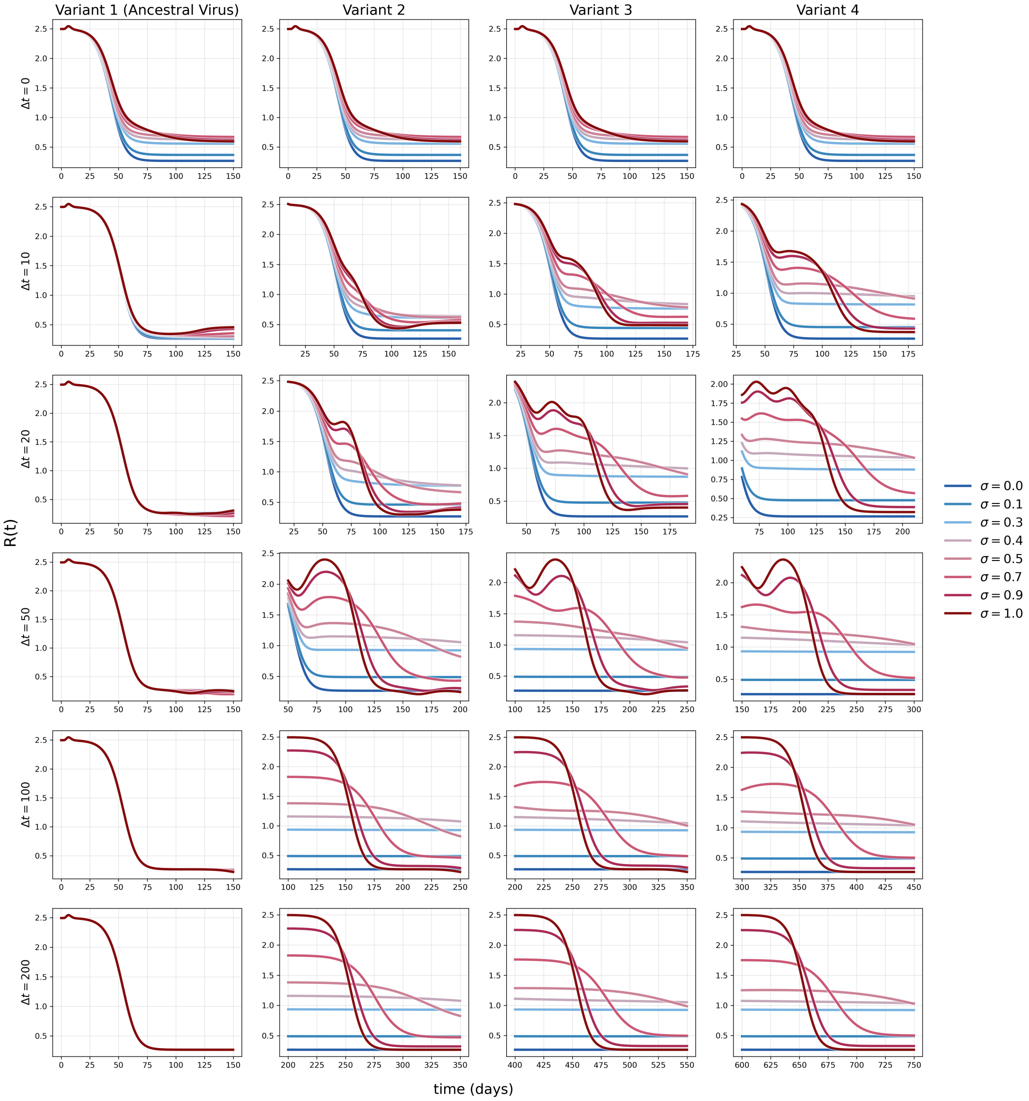

Figure S3: Time-varying reproduction number. Each panel shows the reproduction number as a function of time (days) under different parameter settings. The first to fourth columns correspond to variants 1 to 4, respectively. Each row corresponds to a different emergence delay, $\Delta t = 0, 10, 20, 50, 100, 200$. Different colors indicate different values of the cross-immunity parameter $\sigma$.
