## Supplementary Table of GISAID Accession Numbers and Genome Metadata for "Role of heterogeneous infection histories on variant dynamics: exact and approximate approaches"

### Supplementary Appendix

All genome sequences and associated metadata supporting the findings of this study can be accessed through the persistent digital object identifier https://doi.org/10.55876/gis8.261003md

In addition to the minted DOI, GISAID also communicates the aggregation of GISAID accession numbers (EPI_ISL_IDs) through the corresponding EPI_SET_261003md identifier to facilitate both, the acknowledgment of all data contributors and the direct retrieval of the underlying data from GISAID used in this study.

#### hCoV-19 Virus Data Summary

| GISAID Identifier | Digital Object Identifier | Number of individual viruses | Data Collection range | Number of countries/territories |
|---|---|---|---|---|
| EPI_SET_261003md | https://doi.org/10.55876/gis8.261003md | 697,002 | 2020-01-10 to 2026-09-21 | 1 |
